# Sequencing for a Lifetime: Professional Perspectives on the Value and Feasibility of Lifelong Genomic Medicine

**DOI:** 10.64898/2026.04.29.26352046

**Authors:** Anna C F Lewis, Ingrid A Holm, Adam H Buchanan, Aaron J Goldenberg, Bartha M Knoppers, Amy L McGuire, Robert C Green

**Affiliations:** Harvard Medical School, Boston, MA, USA; Mass General Brigham, Boston, MA, USA; Broad Institute, Boston, MA, USA; Department of Genetics, Boston Children’s Hospital, Boston, MA, USA; Department of Genomic Health, Geisinger, Danville, PA, USA; Case Western Reserve University, School of Medicine, Cleveland, OH, USA; Centre of Genomics and Policy, McGill University, Montreal, Quebec, Canada; Baylor College of Medicine, Center for Medical Ethics and Health Policy, Houston, TX, USA; Ariadne Labs, Boston, MA, USA

**Keywords:** lifelong genomic medicine, genomic newborn screening, health equity, data governance

## Abstract

**Introduction:** Lifelong genomic medicine, in which stored genomic data informs a lifetime of care, is moving from vision toward reality, with dozens of newborn sequencing projects underway worldwide and reanalysis of data already in clinical practice. Professional perspectives on its value and feasibility have not been examined.

**Methods:** We conducted and thematically analyzed semi-structured interviews with 52 US-based professionals, including clinical geneticists, genetic counselors, primary care clinicians, laboratory personnel, and genomic screening implementers. Interviews explored delivery setting, longitudinal clinical relationships, data reanalysis and recontact, consent, data stewardship, privacy, and the pediatric-to-adult transition. Analysis combined deductive domains with inductive coding, with attention to similarities and differences across stakeholder groups.

**Results:** Participants in all five groups saw clinical value in lifelong genomic medicine, citing the relevance of genomic information at multiple life stages, the ability to requery the genome as circumstances change, and the capacity to act on evolving knowledge. A minority contested this value, favoring more targeted testing. Many participants viewed the model as inevitable and described operational precedent for many component activities, from reanalysis of decade-old data and reclassification pipelines to “sequence once, query often” workflows and EHR-integrated alerts. Participants described substantial gaps in governance, spanning delivery setting, consent, data stewardship, recontact, and the pediatric-to-adult transition, alongside continuing operational challenges such as interoperability. No delivery setting was clearly favored: participants saw newborn screening as offering the strongest equity case but as conflicting with the explicit consent it would require, health-system delivery as excluding those without access, and commercial models as potentially short-lived. Participants raised equity concerns at enrollment, in maintaining follow-up, and at the pediatric-to-adult handoff, which we argue are cumulative and mutually reinforcing for those least connected to health systems.

**Conclusion:** Many participants expected lifelong genomic medicine to be implemented, but no comprehensive or clearly favored model for delivering it responsibly emerged. Coordinated investment in governance is needed, alongside evidence on which forms of lifelong genomic medicine offer enough clinical value to justify their costs.

## Introduction

A proposed vision for the future of population-wide genomic medicine is to sequence babies and to keep the genome “on file” to inform a lifetime of care [1–4]. For example, Biesecker et al., advocate for the genome to be sequenced at birth, and genetic results to be sequentially disclosed over time “at a life stage when interventions are known to be beneficial” [1]. They give examples of newborn screening conditions at birth, Wilms tumor and retinoblastoma to children, aortopathy and cardiomyopathy to teenagers, and carrier risks and colon and breast cancer risk to adults. Storing an individual’s genome could enable querying for information that might shed light on a newly developed clinical phenotype. It could also enable recontact when knowledge changes, either because new information changes the classification of variants [5], new genes are linked to conditions [6], or because new interventions — and in particular an increasing array of gene and cell therapies — suggest the benefit of reporting more results. We refer to this vision of sequencing once near birth and returning to the stored data to inform a lifetime of care as “lifelong genomic medicine,” and it has long animated the field of genomic medicine [7]. The term covers the activities involved in reusing genomic data over time, including storage, reanalysis, integration into the electronic health record, and recontact, but not retention of biological samples. Stored data could therefore be queried at set ages, when an individual’s circumstances change, or when knowledge changes. We use “delivery” to refer to the institutional setting responsible for offering sequencing and for the storage, reanalysis, recontact and return of results that follow. With improvements in technology, data management and AI, there is genuine excitement about this finally being possible, though the evidence for the clinical utility of population-scale genomic screening remains inconclusive [8].

Many research projects around the world are sequencing newborns to study the integration of genomic sequencing into public health newborn screening programs (NBS) or the newborn period [9]. The prospect of lifelong genomic medicine is hence becoming less abstract. For example, the UK government is currently sequencing 100,000 newborns as part of Genomics England’s Generation Study. In addition, they aim to “explore the risks and benefits of storing an individual’s genome over their lifetime” [10]. Outside the context of screening newborns, there are several implementation studies that maintain genomic data on file as a resource, for example from the Geno4ME Study [11,12]. Several companies are also offering a sequencing and storage model that becomes a “lifetime digital genomic resource” [13].

It may, however, be that the benefits of keeping a genome on file are outweighed by costs of referring back to legacy data, the risks of discriminatory misuse, and the challenges of data governance [14,15]. Alternative models include not storing the genomic data and instead resequencing the genome if there is a clinical indication [16], or only sequencing a subset of the genome (i.e., panels) [15].

Because many of the component pieces of lifelong genomic medicine are already in place — storage of data sequenced in the context of a clinical indication, reanalysis of genomic data, recontacting patients if variants are reclassified, construction of age-based panels based on actionability — the views of professionals on the feasibility and value of lifelong genomic medicine could refine this vision, including potentially suggesting that the vision should be abandoned. As those who would be responsible for delivering, interpreting, and communicating genomic information across a patient’s lifetime, healthcare professionals have relevant experience of both its potential clinical value and its feasibility, and their views may shape whether such programs are adopted.

Two qualitative interview studies with Australian healthcare professionals on genomic newborn screening touch on the potential for genomic data to be retained and reused across the life course, with participants in both studies seeing value in long-term storage for future reanalysis [17,18]. To our knowledge, no studies have examined professional perspectives on lifelong genomic medicine as a central question.

We conducted semi-structured interviews with over 50 US-based professionals to gain their perspectives on retaining and reusing genomic data across a lifetime, including clinical geneticists, genetic counselors, lab personnel, primary care clinicians (PCCs), and those with experience implementing population screening in healthcare systems. Here we report their perspectives on the value and feasibility of lifelong genomic medicine. The aim was descriptive; we did not set out to determine whether lifelong genomic medicine should be pursued. Results related to the staging of genomic information by age will be reported separately.

## Methods

### Participants

Multiple types of professionals were recruited into this qualitative interview study to enable triangulation of perspectives. This included clinical geneticists, genetic counselors (pediatric, adult, and maternal fetal medicine) and primary care clinicians. While the genetics providers have the most experience with genomic medicine, primary care clinicians would likely be closely or even primarily involved if lifelong genomic medicine were to be implemented. Laboratory personnel, mostly laboratory directors, were also included, as they have experience managing genomic data. Finally, we included those who have led operational roles integrating genomic screening (i.e., genomic testing in the absence of an indication) into health systems. Most were clinically trained (genetic counselors, physicians, or a pharmacist) and held leadership roles in these programs; others included a social and behavioral scientist and a family advocate. We refer to this group as “genomic screening implementers”. All genetic counselors were licensed and most were in clinical practice; several also held research or leadership roles, and one worked primarily as a research genetic counselor. Because participants often held more than one role, each was assigned to the group through which they were recruited.

We used purposive sampling to recruit individuals from all groups. An invitation to participate was distributed via email, some individualized and some via listservs. Primary care clinicians, clinical geneticists, and genetic counselors were recruited mainly through the BabySeq Project sites [19,20] and affiliated institutions described below; laboratory personnel and genomic screening implementers were contacted individually. We used snowball sampling to recruit additional participants, asking interviewees for recommendations of others we should interview within their own institutions, particularly those who might have different perspectives or different subject area expertise on the themes covered. There were no exclusion criteria beyond being a US-based professional in one of these groups.

We recruited healthcare providers from some of the BabySeq Project sites [19,20]. The BabySeq Project studies the integration of genomic sequencing into newborn care and has recruited hundreds of babies across the US. For our study, the BabySeq Project sites we recruited from were Boston Children’s Hospital, the University of Alabama Medical Center in Birmingham, Corewell Health, Michigan, and Children’s Hospital of Philadelphia, including those working in academic hospitals and community clinics. We also invited genetic counselors at the University of Pennsylvania Health System, a sister institution to the Children’s Hospital of Philadelphia. The study was advertised to individuals at these institutions via email listservs. Response rates could not be calculated for participants recruited, as the total number of recipients was not ascertained.

We identified the most active laboratories that conduct genomic sequencing in the US from the list of top submitters of variants to ClinVar and contacted their laboratory directors via email with an invitation to participate [21]. Of 14 emailed, 7 accepted the invitation. For genomic screening implementers (those with experience implementing programs that have launched genomic screening programs), a narrative review identified ten clinical and research programs that use genomic sequencing for population-based screening in a healthcare system in the US [22]. The names and email addresses of individuals associated with these programs were ascertained from publicly available information and the authors’ knowledge. The list was supplemented with others known to the research team to be involved in projects not covered by the review. Email invitations were sent to 18 individuals from this list, 12 of whom agreed to participate.

Practice characteristics were not collected systematically; we describe them from participants’ accounts of their professional practice at the start of each interview and from recruitment records.

All prospective participants received a fact sheet with basic information about the study, including its purpose, before agreeing to participate (see online supplementary material).

### Data Collection

Semi-structured interviews were conducted using an interview guide (see online supplementary material) that was designed based on review of the literature and knowledge of the research team. Some components of the interview guide were common across all types of providers; some were specific to clinical versus operational roles. Topics covered included desirability of lifelong genomic medicine, and facets of current day processes to understand feasibility, including, for example, clinician experiences with longitudinal relationships, current treatment of sequence information after a report is generated, privacy considerations. All participants were introduced to lifelong genomic medicine as sequencing once near birth and querying the data many times to inform care over the lifespan; the activities this would involve were not specified. Where delivery setting had not come up, participants were asked which setting they had imagined (for example, clinical care or newborn screening) and which they thought best. This question was not asked systematically, and participants were not presented with defined delivery settings.

All interviews were conducted by AL between August 2024 and February 2025. Interviews were approximately 60 minutes long, conducted over Zoom videoconferencing and recorded. Only the interviewer and interviewee were present. Field notes were taken immediately after each interview. Auto-generated transcripts were manually cleaned and de-identified to protect privacy and confidentiality. Transcripts were not returned to participants. A short demographic survey was emailed to participants after the interview. Interviewees were mailed a $100 check as a thank you for their time. No repeat interviews were conducted.

### Analysis

De-identified transcripts were coded by AL. Analysis drew on principles of framework analysis [23,24], combining deductive and inductive elements. Domains were defined *a priori* based on the research questions. The study adopted a pragmatist orientation, prioritizing the generation of findings with direct relevance to policy and practice. Within each domain, transcripts were inductively coded using Atlas.ti software using *in vivo* and descriptive codes [25]. Codes were iteratively grouped into themes and subthemes through repeated comparison across codes, transcripts, and stakeholder groups; the coding framework underwent multiple revisions as groupings were refined. Five transcripts, one from each stakeholder group, were coded by a second author (IH) who helped identify themes and refine the codebook. Code frequency was tracked by the five stakeholder groups throughout, enabling systematic comparison of where professional perspectives converged and diverged. Throughout analysis, AL maintained analytical memos documenting evolving interpretations as the coding framework developed. Emerging themes were additionally sense-checked through presentation to and discussion with research colleagues. No substantively new themes emerged in the final interviews within each participant type. Participant checking of findings was not conducted; given the multi-stakeholder design and the analytical focus on cross-group comparison, we prioritized peer debriefing and co-coding over individual participant validation. The complete coding framework, including code frequencies by stakeholder groups, is provided in the online supplementary material. To indicate prevalence, we use “a few” for 2–3 participants, “some” or “several” for 4–8, “many” for 9 or more, and “most” for more than half of those who discussed a topic. Counts for all terms refer to participants who discussed the topic; because not all topics were discussed with all participants, they do not indicate how many participants held or rejected a view. Views described as held “across participant groups” were raised in at least four of the five groups.

During this process, coded materials pertaining to population genomic screening broadly — rather than lifelong genomic medicine — were separated for analysis elsewhere. This paper focuses specifically on lifelong genomic medicine. Codes related to the staging of information by age are also analyzed elsewhere.

As a researcher working at the intersection of genomics and ethics, AL (DPhil, she/her) was a partial insider to the professional community studied. This may have facilitated candor but may also have led some participants to foreground ethical considerations. A small number of participants were known to AL prior to the study, which may have made them more candid or more inclined to give answers they expected the interviewer to share. All participants were informed of the confidentiality of their responses. IH (MD) is a clinical geneticist and researcher, and her expertise provided an additional analytical perspective and helped surface assumptions that might otherwise have gone unexamined. Both AL and IH are experienced qualitative researchers.

This study was approved by the Mass General Brigham IRB, which judged it exempt. The study is reported in accordance with the COREQ checklist (see online supplementary material).

During the preparation of this manuscript, the corresponding author used Claude (Anthropic) to assist with reframing and refining points in the written text. All content was subsequently reviewed and edited by the authors, and the authors take full responsibility for the accuracy and integrity of the published work. Generative AI was not used for data collection, coding, or analysis.

## Results

### Participant characteristics

Fifty-two professionals participated, including genetic counselors (n=16), clinical geneticists (n=6), laboratory personnel (n=8), primary care clinicians (n=10), and genomic screening implementers (n=12). Participants were predominantly female (n=28) and White (n=36), with a mean age of 45 years (range 27–69) and a range of career experience from early-career to senior professionals (Table 1). Participants came from more than 20 organizations spanning all four US Census regions. Fifteen of the 16 genetic counselors and nine of the 10 PCCs were recruited through BabySeq Project sites or affiliated institutions; all seven adult genetic counselors practiced at a single academic health system, most in oncology. Nine of the 10 PCCs cared exclusively for children. Five of the eight laboratory personnel worked in commercial laboratories and three in academic laboratories. The 12 genomic screening implementers each came from a different organization: eight from health-system or academic screening programs, three from genomic sequencing companies, and one from a family advocacy organization. Ten implementers were clinically trained (genetic counselors, physicians, or a pharmacist). Many participants across groups also held research or leadership roles.

**Table 1:**
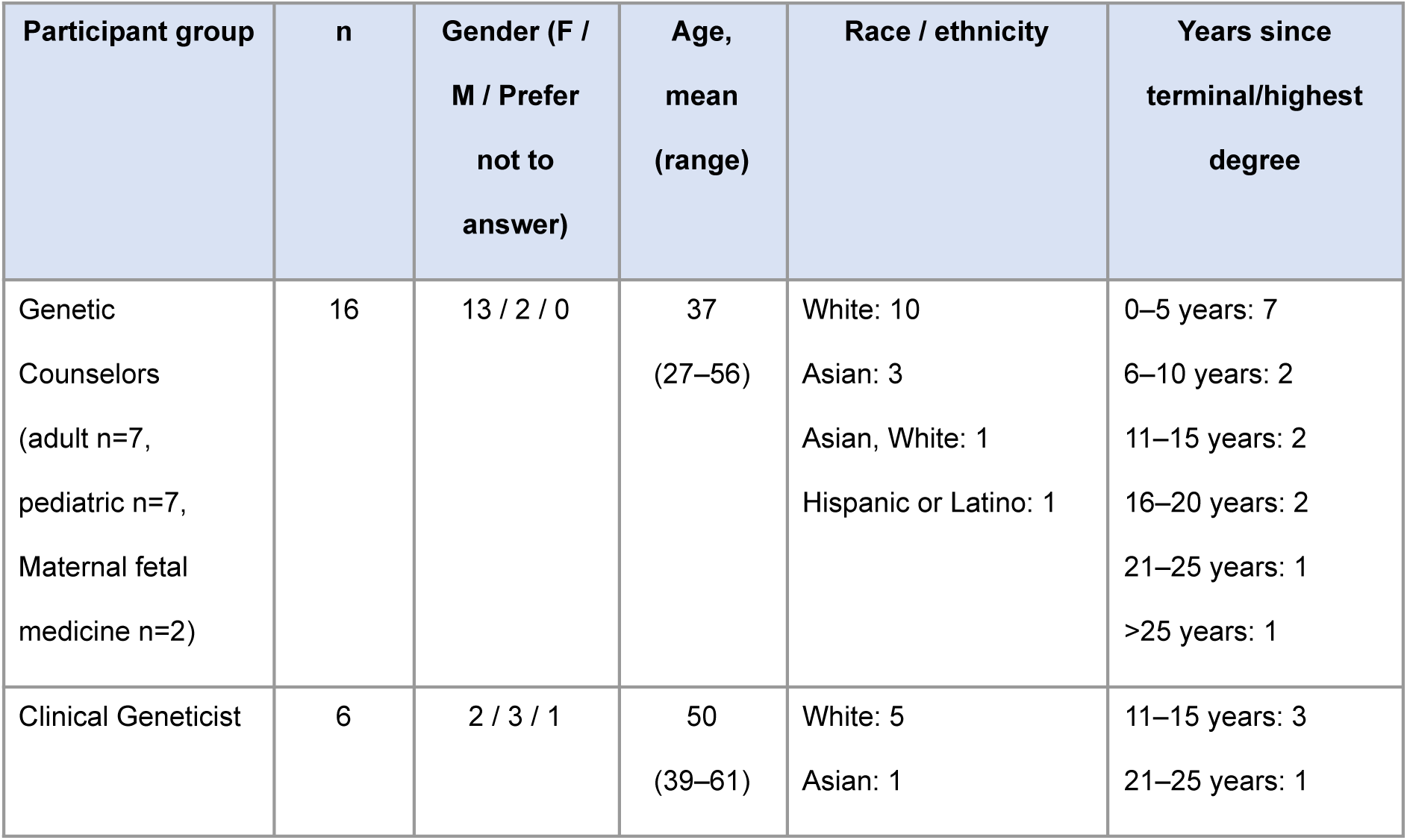
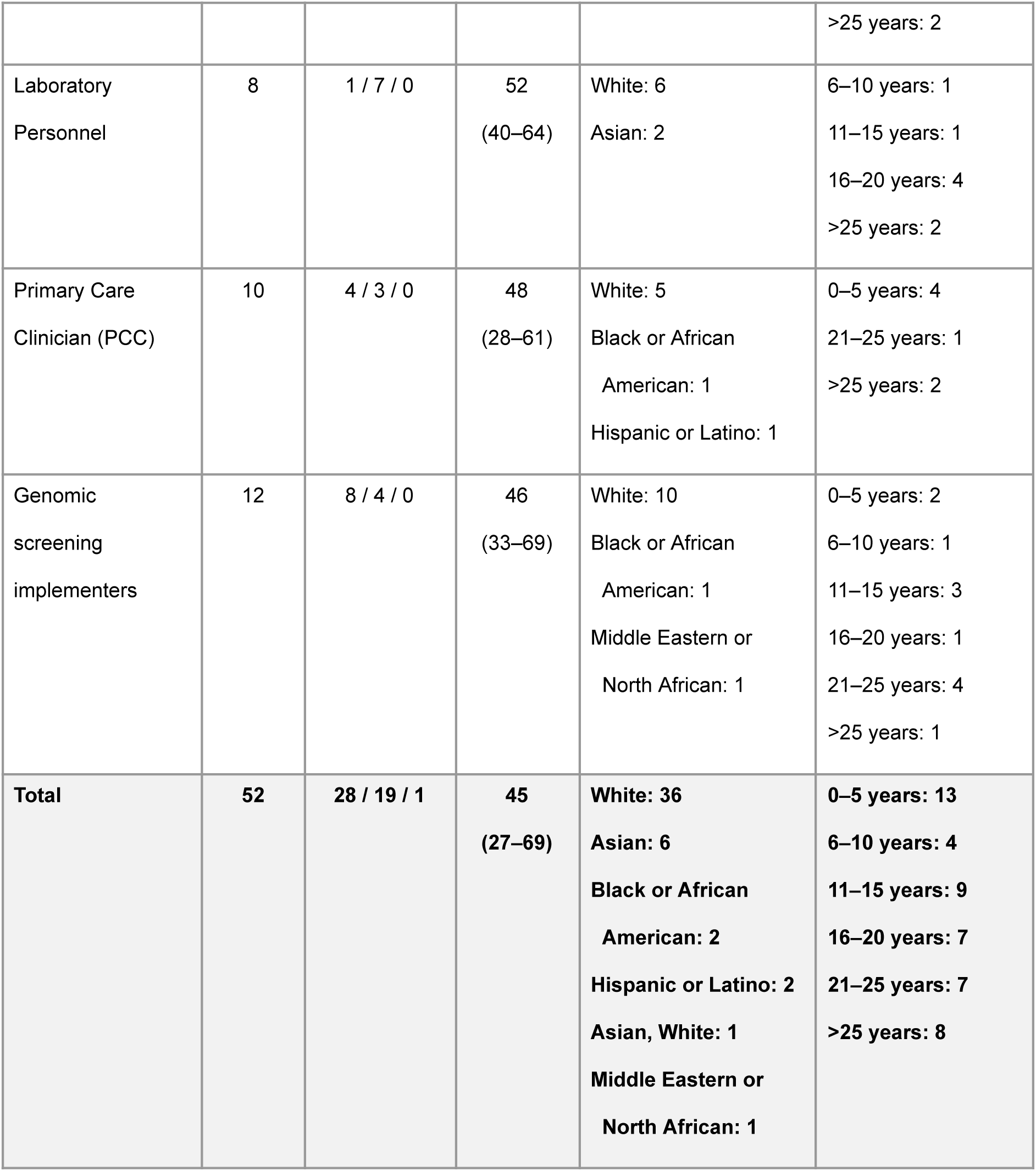
Participant Characteristics. Demographic data were missing for 4 participants (3 Primary Care Clinicians and 1 Genetic Counselor (adult)) and are excluded from the counts presented.

### Perceived value of lifelong genomic medicine

Many participants felt that the lifelong genomic medicine model has clinical value, could help advance future research, and a few saw it as potentially more cost effective than other models of genomic medicine. Although they recognized the complexity of this approach, and some questioned whether the investment needed was justified in light of alternative models that are available, many felt that the lifelong genomic medicine model is already being developed and its implementation is inevitable.

#### Clinical value across the lifespan

There was broad agreement across participant groups that genomic information becomes relevant at different life stages, making the lifelong genomic medicine model clinically and intuitively attractive, see Figure 1. Examples cited included adult-onset actionable conditions, carrier screening at reproductive age, and polygenic risk for common disease, including predispositions relevant to adolescence such as mental health and substance use. (These examples differ considerably in evidence base and actionability; we report them as participants raised them.) Participants, particularly laboratory personnel, also saw value in the lifelong genomic medicine model when individual circumstances change. This included when medications are prescribed (pharmacogenomic results), when family history changes, and when phenotypes emerge or change. Scientific progress was also identified as a source of ongoing value. This included the evolution of knowledge linking genotype to phenotype, both for updates to variant-level pathogenicity classifications and new associations between genes and diseases. It also included updates to guidelines, which could include new interventions associated with particular genetic variation, or new data on optimal screening regimens.

**Figure 1:**
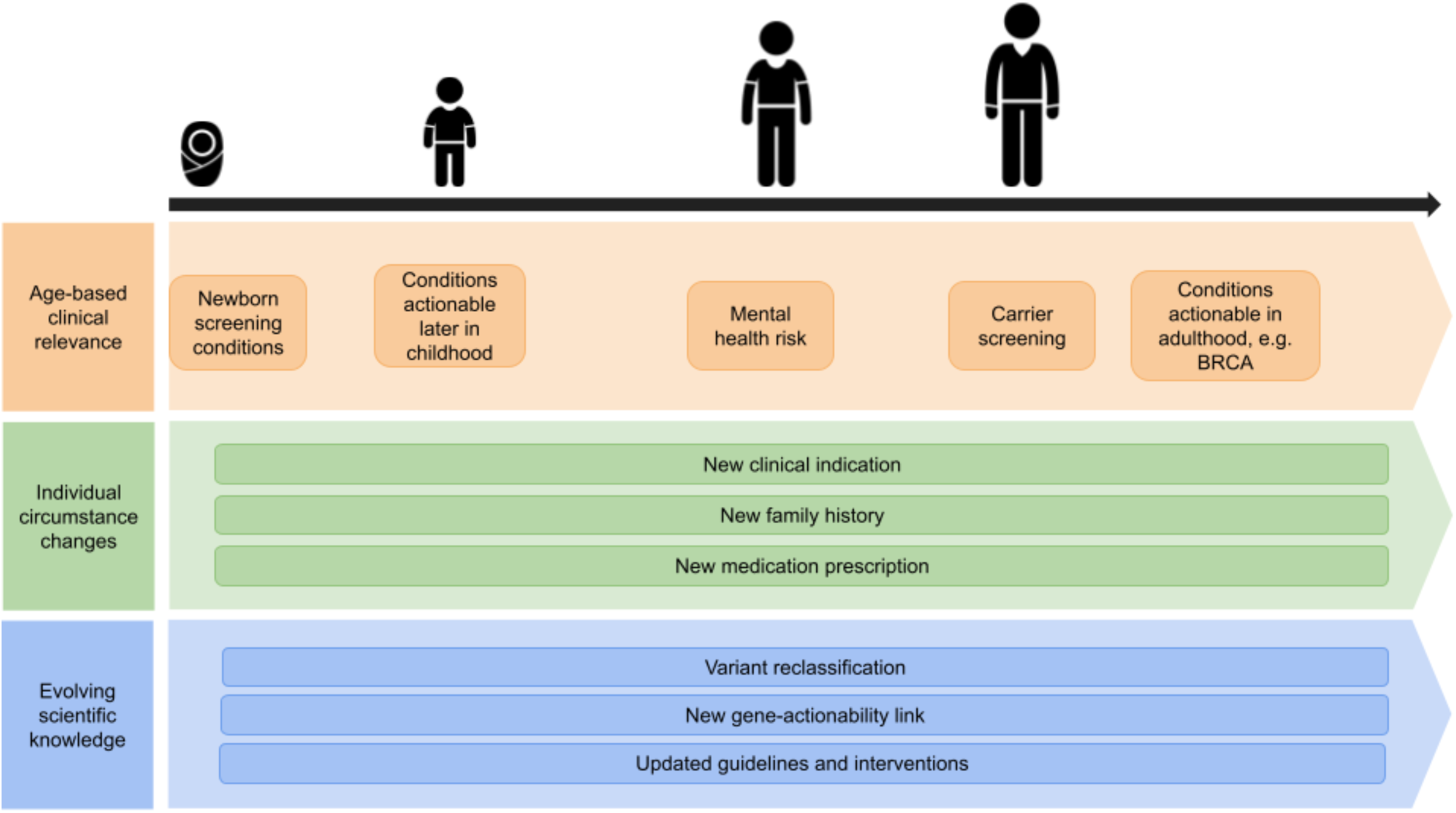
Clinical value of the lifelong genomic medicine model.

Beyond individual clinical benefit, participants, in particular genomic screening implementers, identified the potential for broader population-level learning to advance research, improve variant interpretation, and enable a learning health system. Moreover, this value, which would require appropriate consents to be in place, could aid institutional buy-in.

Participants saw value in storing and reanalyzing genomic data over retesting. Genetic counselors and lab personnel noted that compared to retesting, reanalysis was cheaper, faster, and does not require a new sample. Others stressed that current practice of not reusing data is wasteful. As one genomic screening implementer put it, *“if you think about how we use genetics traditionally, it’s a waste… You order these multiple tests, they land in the medical record, they get lost, people get multiple tests ordered of the same thing”* (P24, genomic screening implementers).

The lifelong genomic medicine model depends on having broad genomic data available for future queries, and genetic counselors and genomic screening implementers saw this as justified because, in their experience, broader testing had repeatedly proven its value. They also noted that the increasing norm of running virtual panels on exome or genome backbones meant the distinction between “panel” and “genome” was becoming artificial. One genomic screening implementer noted that patients themselves sometimes expressed frustration that previously collected genomic data could not be re-analyzed.

#### Perceived inevitability and complexity

Across many participant groups there was a pervasive sense that some form of lifelong genomic medicine was inevitable. For example one pediatric genetic counselor noted *“I have yet to see any major barriers that would stop this train from going down the tracks”* (P3, GC pediatrics). Genomic screening implementers were particularly emphatic, for example *“every other system that we talk to, with very few exceptions, are bullish. They’re like, ‘Yeah, this isn’t an if, it’s a when’”* (P24, genomic screening implementers).

Participants across most groups, particularly genetic counselors, articulated the complexity involved in implementing the lifelong genomic medicine vision. As one adult genetic counselor observed, *“I mean, it’s not impossible. I just — there’s so many moving parts”* (P43, GC adult). Another participant articulated concern about reputational risk if the complexity were not adequately handled. Genomic screening implementers counterbalanced these concerns with pragmatism: it is acceptable not to get everything right the first time, and that the scale of the challenge should not be allowed to freeze the process.

#### Questioning the value of lifelong genomic medicine

A few participants questioned whether investment needed for lifelong genomic medicine was justified. The argument that finite resources might be better directed toward expanding access to interventions with a more established evidence base — what a few described as “low-hanging fruit” — was made most pointedly by one laboratory participant, but also by a few genetic counselors and genomic screening implementers. One lab participant captured the counter narrative to data accumulation: *“I feel like my opinion has changed dramatically over the last decade. I very much began as a ‘data is awesome, more data is more awesome’ person, and now I’m more in the camp of ‘data is a burden’. There are important questions that you can answer effectively now, and there’s ways of making that harder for yourself.’ In many cases, more data just makes it harder to do the thing that you’re trying to do now”* (P10, lab personnel).

Some participants raised retesting rather than reanalysis as an alternative to maintaining stored data. One clinical geneticist noted that it was currently faster to order a new test than to get reanalysis results. And two participants described the “one and done” philosophy as naive. *“I’m wary of a public endeavour to say just have this one test, and you’re set up for life, because that’s not the truth… There’s no one single diagnostic test that’s going to cover you for everything. I talked to so many parents about this, and they say, ‘So I never have to have any other genetic test, because I had all my genetics looked at?’”* (P3, GC pediatrics).

Two participants, a laboratory participant and a PCC, questioned the need for whole genome sequencing. They stressed that genome-wide data is very complicated, and that there is no other area of medicine where we collect information because it might be useful in the future, and that they believe the focus on collecting genome-wide data drives payer anxiety about downstream costs. An advantage of gene panels raised by one primary care clinician (PCC) is that panel testing could remove the burden of choice from parents about what to learn when.

### Perceived feasibility of lifelong genomic medicine

Participants identified a series of structural prerequisites for lifelong genomic medicine, spanning the choices of delivery setting, data governance, patient-facing processes, the longevity and reusability of stored data, and the mechanisms by which stored data would generate clinical action over time. For many individual components, operational precedent exists; the challenges lie in connecting them into a coherent system, and in governance gaps that cut across nearly every area. Delivery settings are summarized in Table 2. Figure 2 maps the component infrastructure required; Table 3 details the operational precedent, unresolved challenges, and equity implications for each.

**Figure 2:**
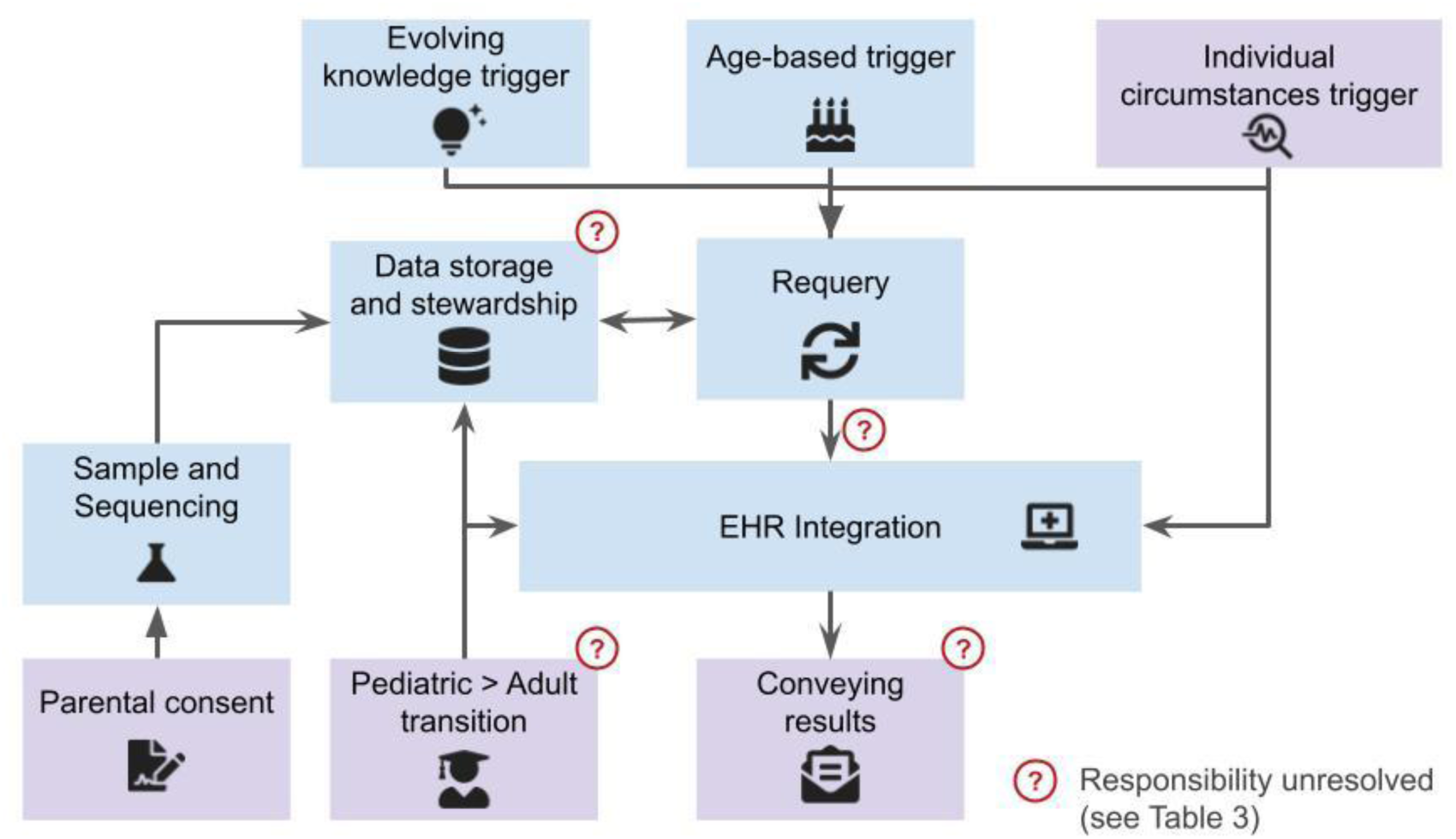
Schematic of the component infrastructure required for lifelong genomic medicine. Boxes represent functional components, and arrows indicate dependencies and data flow between components. Purple boxes represent patient-facing processes. Requery refers to reanalysis of stored data. Question marks indicate points where participants described responsibility as unresolved. Operational precedent, unresolved challenges, and equity implications for each component are detailed in Table 3.

**Table 2:** Delivery settings for lifelong genomic medicine, as discussed by participants. Cells summarize participants’ views of each setting. Settings are not mutually exclusive; some participants suggested a staged approach beginning in clinical settings.

|  | **Newborn screening (NBS)** | **Health system / Clinical** | **Industry / Direct-to-consumer** |
| --- | --- | --- | --- |
| **Equity of reach** | Strongest equity case: reaches nearly every child. But NBS not always equitable in practice | Excludes those without access to participating health systems. Currently a boutique, out-of-pocket product |  |
| **Consent model** | Fundamental incompatibility: NBS currently operates under implied consent; lifelong genomic medicine would require explicit consent. Risk of destabilizing public acceptance of existing NBS | Consent can be embedded in clinical relationship (e.g. integrated into annual physical with e-consent and education) | No data on consent models in this context |
| **Funding / sustainability** | State NBS programs drastically under-resourced for genomic integration | Revenue model unresolved: unclear if initial cost includes revisiting, storage, or unmasking. Some health systems investing upfront (nominal fee, then bill insurance for phenotype follow-up). Depends on convincing payors of economics | Challenging business model |
| **Data stewardship** | Not designed for non-infant-relevant information | Health system as steward, ideally hospital affiliated with primary care clinic. But competing interests in data control (AI, commercial uses); one participant would not trust health systems as steward | Data can persist outside health systems. But companies may not last: three participants independently raised bankruptcy risk; patients expect data to remain usable when companies fold |
| **Durability** | State programs are permanent infrastructure if resourced | Patients change health systems; data does not follow. Health systems want to control the whole process but this is not feasible | Least durable: companies may go under; relationship with testing company may not be lifelong |
| **Scalability / readiness** | Existing infrastructure to build on; cross-correlation with biochemical NBS possible; advantage of uniformity within a jurisdiction. But needs substantial provider and public education first; turnaround time longer for NGS than NBS | Already happening in some settings (e.g. concierge healthy whole genome, integration into annual physical). But not at population scale | PCC-support model proposed (company supports PCC, PCC supports patient). But limited discussion overall |

**Table 3:**
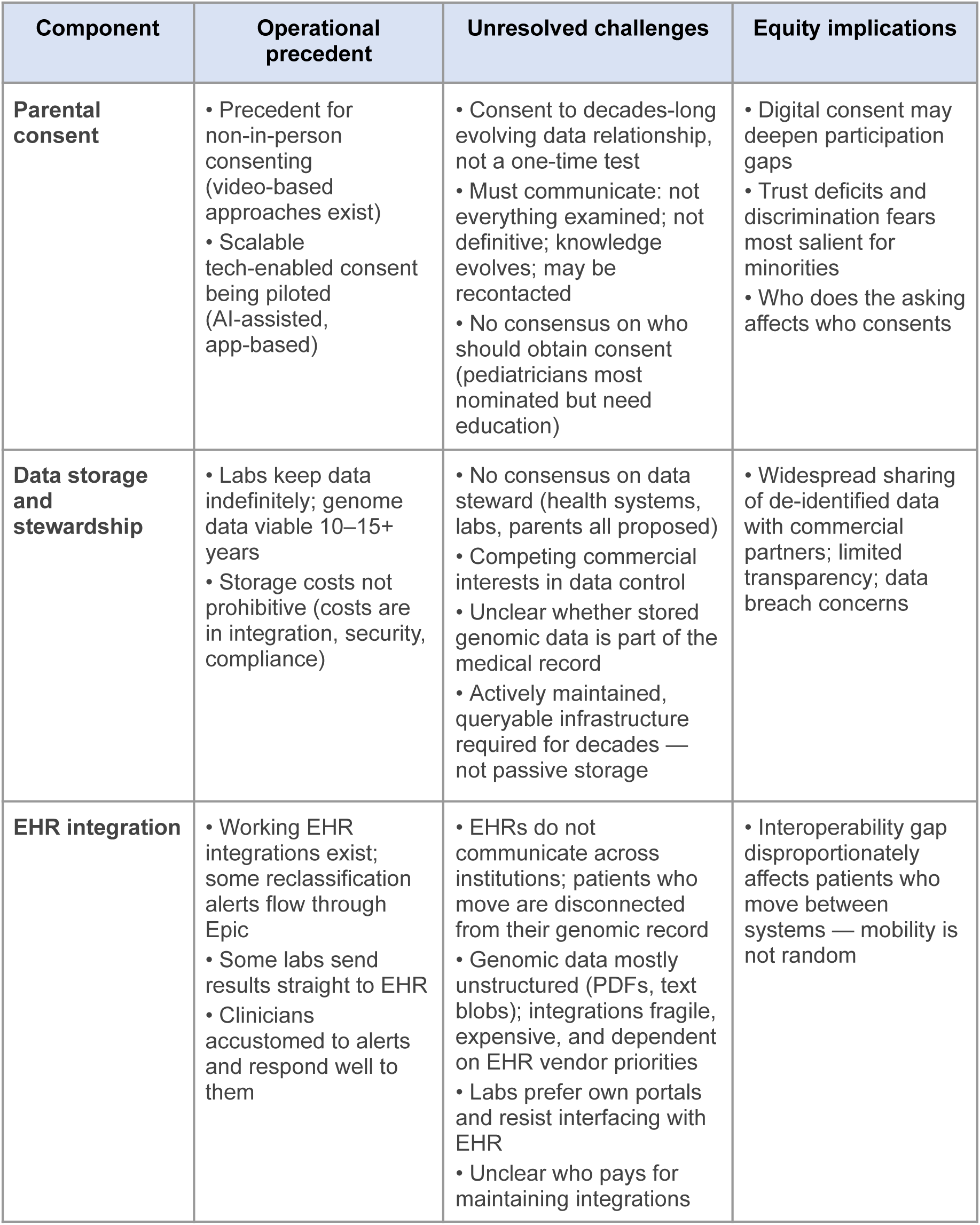
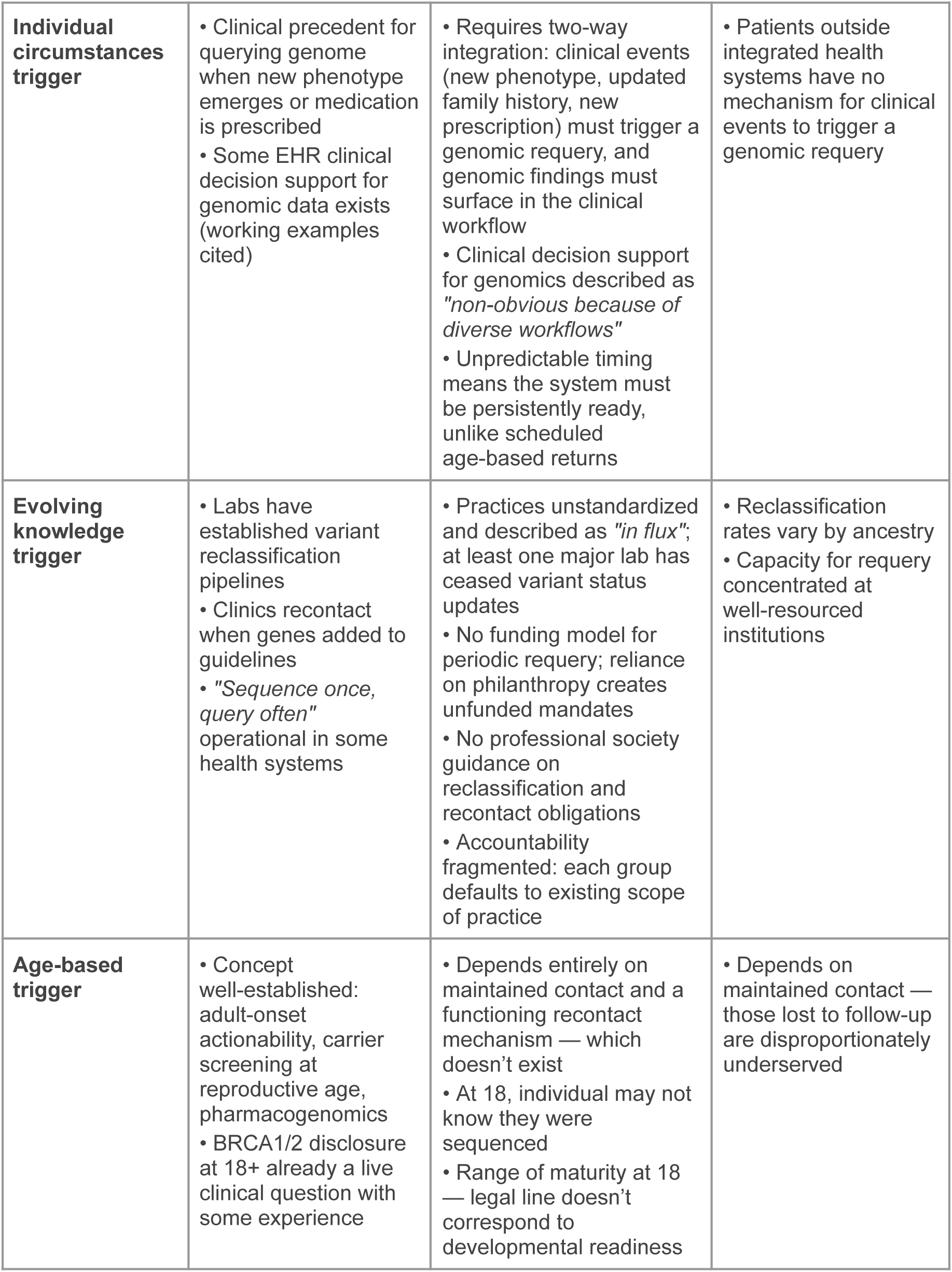
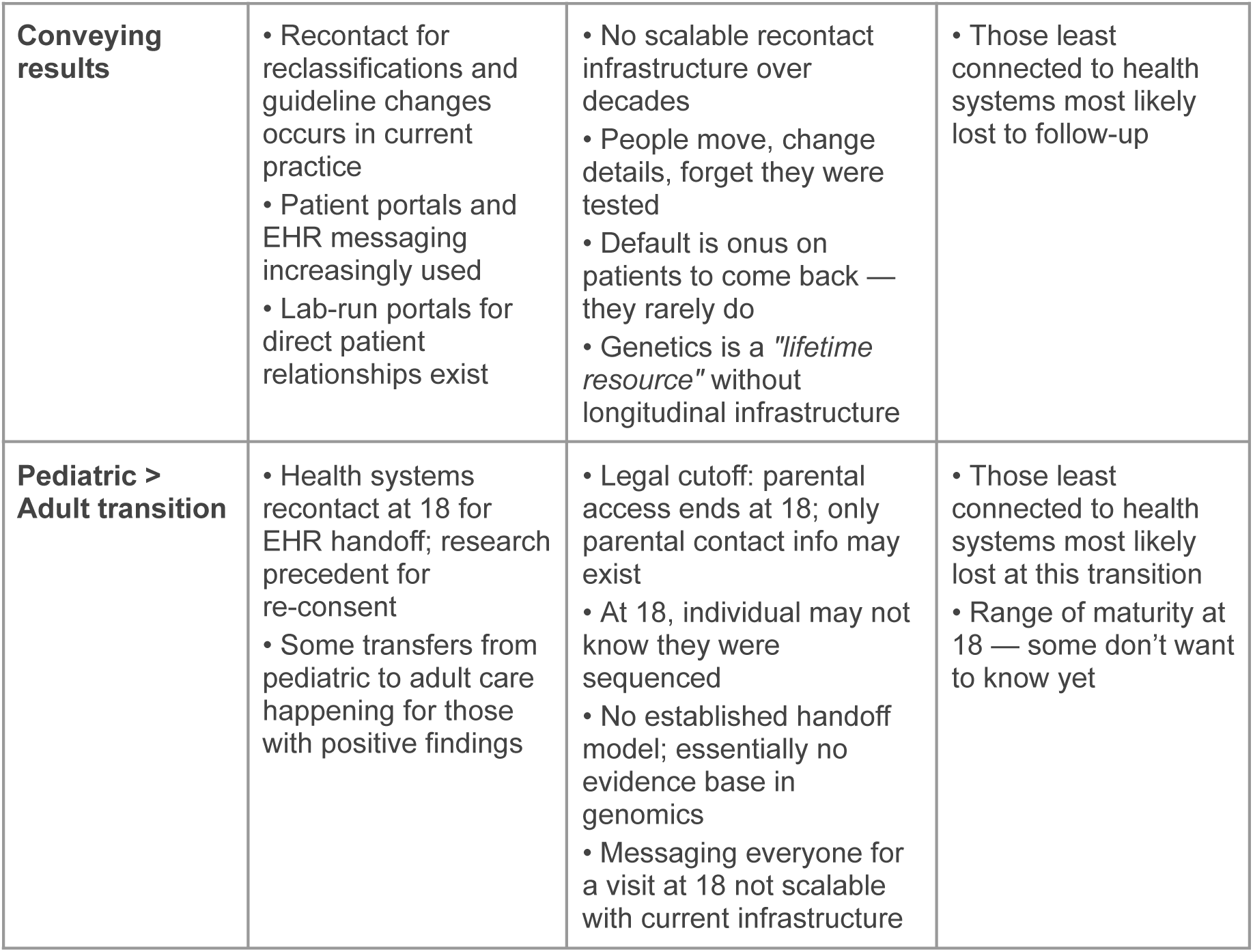
Feasibility landscape for lifelong genomic medicine. For each component of the envisioned model, operational precedent, unresolved challenges, and equity implications are identified from participant interviews. Components are ordered to reflect the flow from initial sequencing through longitudinal management.

#### No clear delivery setting

Participants who discussed delivery raised three possible institutional settings and did not agree on which was best placed to deliver lifelong genomic medicine (Table 2). Questions about revenue models and long-term financial sustainability arose repeatedly. Who would pay for the initial sequencing, subsequent reanalyses, and data storage is unresolved, and it isn’t clear if insurance would cover those costs. A few participants acknowledged that there is no obvious access pathway for lifelong genomic medicine in the US, given the disseminated nature of healthcare in the US — a difficulty it shares with other preventive services.

Health systems were identified by a few, mainly genomic screening implementers, as the most natural home for such a program, given their ability to coordinate longitudinal care, integrate data into electronic health records (EHRs), and maintain patient relationships over time. A few genomic screening implementers noted that healthy whole-genome offerings were already beginning to emerge as clinical products. However, participants also recognized that health systems do not serve all patients equally, raising equity concerns, including for families far from specialist services: *“Transportation is the rate-limiting step to health. I live 2 hours from [city], in a very rural area with spotty Internet. And now you’re going to tell me that my kid has a mutation of some kind, and then I have to meet with the genetic counselor in [city], either on Zoom or in 6 months. I’m good. My baby looks fine to me”* (P31, primary care clinician).

Some participants advocated for the integration of lifelong genomic medicine into NBS, primarily on the basis of equity arguments, as NBS reaches almost every child. Other advantages noted included the ability to cross-correlate with biochemical NBS, the uniformity that would be possible within a jurisdiction, and the existence of a pre-existing model. Arguments for separating lifelong genomic medicine from NBS included: the fundamentally different consent regimes that would be appropriate for lifelong genomic medicine compared to NBS, which currently operates mostly under implied consent; the lack of staffing or funding within state NBS programs; the fact that NBS is not designed for non-infant-relevant information; and the need to protect existing NBS programs from destabilization. Some participants, among laboratory personnel, primary care clinicians, and genomic screening implementers, suggested lifelong genomic medicine could start in the clinical context, and migrate to NBS as infrastructure improves, evidence accumulates, and acceptance matures.

A few genomic screening implementers identified roles for industry partners, including supporting PCCs and providing data persistence outside health systems through direct-to-consumer channels, though clinical geneticists and lab personnel questioned the durability and business viability of commercial partners.

#### Patient- and parent-facing processes

With lifelong genomic medicine parents would be consenting not to a single test but to an open-ended ongoing relationship, concerning data whose clinical significance will change. Parents would need to understand that not everything will be examined, that knowledge evolves, and that they may be recontacted. Participants across several groups suggested that consent at this scale would need to be delivered through scalable approaches, including video-based and AI-assisted consent (Table 3).

Privacy concerns were present across participant groups but were not dominant; several described the proportion of patients who actively objected to genomic data storage as small. However, the indefinite timeframe of the lifelong genomic medicine model compounds these concerns in ways that are distinctive to this model. Participants described commercial data sharing practices as already widespread but poorly transparent, and high-profile breaches such as 23andMe had shaped patient expectations about data security. More prominently, fear of insurance discrimination persisted regardless of existing legal protections and was raised across participant groups as a primary driver of whether patients would consent.

Participants also identified mistrust of institutions, particularly among minority communities, as shaping who would engage with a program that asks for open-ended data retention; one primary care clinician described “way more distrust” of newborn screening among the minority families they served (P32, primary care clinician).

Managing the transition from pediatric to adult care was identified as a structural vulnerability of the lifelong genomic medicine model, discussed across participant groups and with particular depth by genetic counselors and clinical geneticists (who also pointed out that this transition is challenging across all of medicine). There is some precedent to rely on, but concerns about scalability were common. One genomic screening implementer described their plan: *“we’re going to start sending them a MyChart message that says… you’re 18. Now you have ownership of your genomic data. Would you like to come in and sit down with us… in the short term, that’s our plan, but I don’t know how we do it in the long term”* (P29, genomic screening implementers). The challenge is not only logistical but legal, with parents losing access to a child’s medical record when the child turns 18: *“The kid turns 18, we’re not allowed to contact the parents anymore now, but we only have the parents’ contact information. I guess the logical thing would be to contact the parents at 17 years, 11 months, or something. None of this is very well structured, actually not structured at all yet”* (P2, clinical genetics). Within genetics, the field is in the early days of working out how to manage this handoff, with current practices varying, for example regarding whether reanalysis continues after the child turns 18 years. Some participants framed the transition not only as a vulnerability but as an opportunity to reengage the individuals sequenced.

Sustaining contact with patients over a lifetime was described as one of the most intractable practical challenges of the model, with broad uncertainty about how this would work in practice. People move, change contact details, and frequently do not update their healthcare providers. People may forget testing even happened. As one clinical geneticist noted: *“Healthcare [is] so screwed up, we can barely take care of the patients we have in front of us, let alone trying to shepherd these patients all throughout their lifetime, and having the ability to give them information back and forth… and none of that coordination generates any revenue for my health system”* (P48, clinical genetics). One pediatric genetic counselor noted that genetics professionals tell patients they are a lifetime resource, while a PCC observed that genetics does not, in practice, maintain longitudinal relationships with patients.

#### Data longevity and reuse

Whether stored genomic data would remain technically viable for reanalysis over the multi-decade timeframe envisioned for lifelong genomic medicine prompted substantive discussion, particularly among laboratory personnel, genomic screening implementers and clinical geneticists. The general view was that whole-genome short-read data was robust and would retain analytical value for at least ten to fifteen years (longer than exome data, which was seen as less future-proof). Laboratory personnel described using data ten or more years old in current reanalysis workflows, and one genomic screening implementer cited examples of disclosed findings derived from sequencing performed over a decade earlier. Other participants had been told by laboratories that older exomes were too outdated to reanalyze, or would prefer to resequence. The transition to long-read sequencing was anticipated as a likely inflection point that would require some reassessment of whether reanalysis or retesting was more appropriate for older samples. But while newer technology might be preferred for individuals with unsolved phenotypes, for healthy individuals enrolled in population screening, existing data was likely to remain sufficient for much longer. One genomic screening implementer expressed the view that the reanalysis-versus-retest decision should not be treated as a barrier to establishing the lifelong genomic medicine model now, noting that the field could address this question as the technology evolved. One lab participant also raised the possibility of retaining the biological sample itself as a complement to or hedge against data obsolescence.

Lifelong genomic medicine requires not just storing data but maintaining a live, queryable infrastructure over decades to inform clinical care. A clear picture emerged from laboratory personnel that whereas data storage costs were not prohibitive, and indeed many labs keep data indefinitely, the significant costs are maintaining integrations between laboratory systems and EHRs, security audits, meeting the regulatory requirements that attach to holding data for long periods, and the infrastructure required to enable authorized access over time. Storage and querying themselves were described as inexpensive; one laboratory participant identified recall of patients, rather than storage, as the costly step.

Data stewardship was identified as an open question by clinical geneticists, lab personnel, pediatric genetic counselors, and genomic screening implementers, with proposed data stewards ranging from health systems, to labs, to parents. A few noted that everyone wants to control data, partly to enable AI applications, and partly because control of data confers commercial leverage. *“[E]verybody’s angling for data storage because they want the data, they want to be able to use it, sell it, research. … [it’s] a business marketplace more than being very patient centric”* (P48, clinical genetics).

#### From stored data to clinical action

While variant reclassification and evolving guidelines affect all genetic testing, lifelong genomic medicine transforms these from incidental possibilities into structural commitments. The value proposition of maintaining genomic data across a lifetime rests on the promise that the data will be revisited as knowledge changes, yet responsibility for acting on that evolving knowledge is fragmented. Participants across several groups, predominantly genetic counselors, described encouraging patients to check back periodically — telling patients that contact is “a two-way street,” or advising parents to return in ten to twenty years to see if knowledge had changed — while acknowledging that patients told to follow up with genetics “never do.” This was recognized as inadequate, but with no clear alternative established. One primary care clinician observed that genetics is “pretty unique in the changing knowledgebase and revisiting data,” reinforcing that this is not a routine challenge with established precedent from other areas of medicine. One genetic counselor noted that liability concerns drove clinical practice in ways not always aligned with patient-centered goals, and a few expressed distress at the prospect of holding potentially actionable information without a clear mechanism to act: *“if we have this information, you wanna make sure you can always get it to those people… we never want to be losing sleep because we have this information, and we can’t get it to the person who needs it” (P28, GC Pediatrics)*.

Participants described substantial operational precedent for the component activities that would underlie this chain. Laboratories had established variant reclassification pipelines and proactively sent updated reports to ordering providers. Genetic counselors described clinics recontacting patients when genes were added to guidelines or panels grew, and some had gone back to research participants when the American College of Medical Genetics and Genomics (ACMG) secondary findings list was updated. Some laboratories had developed streamlined processes for variant reinterpretation, and a few genomic screening implementers described existing “sequence once, query often” workflows in which reanalysis of screening data for new indications was already occurring. However, the current reclassification landscape was described as “in flux,” with practices unstandardized: some laboratories reported only upgrades to pathogenic, others reported all changes, some did not issue downgrades, and reclassified variants did not always result in a new report. Not all updated results were wanted by physicians. At least one major commercial laboratory had ceased providing variant status updates. Current processes were not scalable to population level, with staffing insufficient for recontact when classifications change. Participants recognized that for lifelong genomic medicine to be sustainable at scale, the processes of reanalysis and recontact would need to be substantially more automated, and that there was no funding model for periodic reanalysis, with current reliance on philanthropy creating unfunded mandates that become unmet obligations.

The aspiration across participant groups was that any treating provider would have access to the genome, with relevant findings surfaced through clinical decision support. The lifelong genomic medicine model places specific demands on EHR integration beyond what one-time testing requires: a new phenotype or updated family history would need to trigger a relook at the genome, evolving guidelines would need to surface updated recommendations, and critically, when patients move between health systems their genomic record would need to follow. Participants across multiple groups, particularly clinical geneticists, noted that EHRs in the US do not talk to each other, and that patients who move are disconnected from their data. Working examples of EHR integration for genomic data existed, and two participants described reinterpretations flowing into the EHR. But the current state was described as minimal with integrations fragile, expensive to maintain, and dependent on EHR vendor priorities. Clinical decision support for genomics was described as non-obvious because of diverse workflows. Laboratories were described as preferring their own portals and resisting interfacing with EHRs, creating an unclear boundary between the testing laboratory and the health system.

## Discussion

### Summary of findings

This is the first study to examine professional perspectives on lifelong genomic medicine as a primary research question, drawing on interviews with 52 participants across multiple stakeholder groups. Participants in all groups saw clinical value in maintaining genomic data across a lifetime, and many described lifelong genomic medicine as inevitable. In our view, this reflected expectations about the field’s direction more than evidence of readiness. Much of what lifelong genomic medicine envisions is already happening in fragmented form in different settings, including recontact post variant reclassification [5,26], reanalysis when phenotype changes [6], and population screening programs that maintain genomic data as a queryable resource [11,12]. Participants raised two sets of unresolved questions. Most focused on governance: who is responsible for acting on evolving knowledge, who pays, who maintains contact, and who holds the data. A minority questioned whether the clinical value of lifelong genomic medicine justifies its costs.

### Uncertain and uneven value

The value of lifelong genomic medicine is likely to differ across its component activities. Querying stored data when a new phenotype emerges is an extension of diagnostic reanalysis, which is already established practice with a documented yield [6]. Recontact when knowledge changes also has some clinical precedent and professional guidance [26], although participants described current practice as unstandardized. Returning results by age to people sequenced as healthy newborns has the weakest evidence base: many variants are less penetrant in unselected populations than in clinical cohorts [27,28], although penetrance estimates depend on how clinical features are ascertained and may be higher than code-based estimates suggest [29]. What matters for age-based return is the absolute risk conferred, weighed against the burden of the recommended intervention, and the benefits and harms of population genomic screening at scale remain uncertain [8]. Participants’ view that lifelong genomic medicine has clinical value may not apply equally to each of these activities, and our interviews did not ask them to distinguish between them.

A few participants, most pointedly one laboratory participant, questioned whether the investment is justified when interventions with a stronger evidence base remain underused. One described moving from seeing more data as better to seeing data as a burden that makes present-day questions harder to answer. This view challenges the assumption that storing more data creates more value, and it bears on each governance question discussed below, since every commitment to reanalyze, recontact, or steward data has costs that clinical benefit must justify. The sense of inevitability was strongest among genomic screening implementers, who have both the most direct experience of the component activities and, in some cases, an interest in their expansion. Either could explain their confidence, and on both, a sense of inevitability may make such questions harder to raise. If lifelong genomic medicine is seen as inevitable, both its evidence base and its governance risk being treated as downstream concerns rather than prerequisites for implementation.

### Governance, particularly in the US context

Participants generally regarded long-term data storage as technically manageable, though they noted continuing challenges with changing sequencing technology, interoperability, and EHR integration. They described larger gaps in governance: who is responsible for acting on evolving knowledge, who pays for reanalysis, who maintains contact, who holds the data, who bears liability. Several of these are questions about money rather than technical capacity: coordination, recontact and reanalysis are largely unreimbursed, so the work is feasible but unfunded. Previous work in the context of Australian genomic NBS identified the need for data governance and surfaced the need for consideration of managing the genomic data generated [17,18]. Our study identifies in detail the specific accountability, recontact, consent, and data stewardship challenges that emerge when genomic information is maintained as an evolving resource over decades. Participants also described anxiety over liability and distress at holding potentially actionable information without a way to act on it, echoing findings on recontact in UK clinical practice [30], and expected this burden to grow with the volume of stored data.

Many of these gaps reflect the structure of US healthcare. Insurance coverage changes over a person’s life, records do not follow patients between health systems, and insurers’ coverage rules can conflict with the aims of precision medicine [31]. Participants described these features directly, noting that there is no obvious access pathway in the US and that patients who move are disconnected from their data. Countries with national health systems and national genomics programs, such as the UK’s Generation Study [10], may be better placed to maintain contact and link genomic data to a lifelong health record [32], but would still face the questions of clinical value, consent, and accountability described here.

Participants did not agree on a delivery setting. They saw the near-universal reach of NBS as its main equity advantage, but this rests on its current position as part of public health, whereas lifelong genomic medicine would also depend on the medical health system for follow-up care. Parents already view genomic sequencing as requiring a higher standard of consent than standard NBS [33], and lifelong genomic medicine would add consent to an open-ended relationship with data whose significance will change unpredictably over decades. NBS is also not a single system: state programs differ in the conditions screened, follow-up services, financing, and retention of blood spots [34], so integration would proceed differently across states. Hybrid models already exist, such as North Carolina’s Early Check, which offers voluntary genome sequencing, with parental consent, of samples collected through state NBS [35]. Hybrid models, or the staged approach some participants suggested, beginning in clinical settings and migrating toward NBS as infrastructure and evidence mature, may offer pragmatic paths forward.

Existing frameworks for responsible genomic data governance have been developed primarily for research data sharing contexts [36,37], and professional guidance on recontact after reclassification has been published [26]. Participants did not identify comprehensive, consistently implemented governance arrangements for clinical genomic data held over decades. Proposed stewards ranged from health systems to laboratories to parents, with participants noting that competing institutional interests, including data’s commercial and AI value, risk creating a marketplace dynamic rather than patient-centered governance. The concern about commercial data interests is consistent with public attitudes research [38,39]. Stewardship also has costs: laboratory personnel described the expense of maintaining integrations, security, and regulatory compliance, which returns to the question of whether the value of stored data justifies them.

### Pediatric-to-adult transition

Participants identified the pediatric-to-adult transition as a structural vulnerability. Transition programs are being developed for some genetic conditions, such as hereditary cancer predisposition [40,41], but to our knowledge the transition has not been studied for individuals sequenced as healthy newborns, who may not know they were sequenced. Yet it is when an individual becomes an adult that they could consent for much relevant information, including carrier status and adult actionability conditions. Moreover, in keeping with evolving assent models, the child should be involved with decisions about the return of genomic information, for example during adolescence [42].

### Equity concerns

Participants raised equity concerns about consent, loss to follow-up, and the pediatric-to-adult transition (Table 3), in some cases in response to a direct question about equity. Few of these barriers are unique to genomics; they follow familiar patterns of compounding disadvantage in US healthcare [43]. What lifelong genomic medicine adds is a requirement for contact and follow-up sustained over decades, so in our view the concerns are cumulative and mutually reinforcing: those least connected to health systems are most likely to face digital consent barriers at enrollment, most likely to be lost to follow-up when they move, and most likely to miss the pediatric-to-adult transition handoff. These add to known patterns in which those least well represented in genomic databases are least well served by genomic medicine [44,45]. In a largely private system, lifelong genomic medicine could also widen gaps if people with better insurance are more able to obtain sequencing, reanalysis, and the preventive care that results indicate [31], and if new work for currently healthy people draws on clinical capacity needed by those who are already sick. In our view, equity will need to be designed into lifelong genomic medicine programs from the start.

### Future directions

Several research priorities emerge from our findings. Evidence is needed on the clinical utility and costs of each component of lifelong genomic medicine, particularly age-based return of results to people sequenced without a clinical indication. This study captures professional perspectives; engagement with diverse patient and advocate stakeholders is now needed, including populations most likely to be excluded or to decline participation in lifelong genomic medicine. A scoping review of the reclassification and recontact literature argues specifically for deliberative methods — in which participants discuss, learn from others, and refine their views — as more productive than further surveys within homogeneous samples [46]. Pilot programs specifically designed around the pediatric-to-adult transition in genomic medicine, which participants identified as a structural vulnerability, are a priority. Comparative evaluation of who is reached and who is missed under different delivery settings would inform more inclusive program design. Empirical work on recontact and engagement over time is needed to move beyond speculation about how longitudinal genomic relationships would function in practice.

### Limitations

This study has several limitations. All interviews were conducted and primarily coded by a single researcher (AL), which may have introduced interpretive consistency at the cost of analytical breadth. This was mitigated through co-coding by a second researcher (IH), iterative discussion of the coding framework, and sense-checking of emerging themes with research colleagues. Our sample was US-based, and many of the governance and infrastructure challenges we identify — particularly EHR fragmentation, the absence of a centralized screening infrastructure, and uncertainty about institutional home — reflect structural features of the US health system. Findings may not generalize to contexts with more centralized delivery, though the core questions of accountability, consent, and data stewardship are likely to arise in any setting. We also focused on sequencing near birth, although many of the issues we report would apply to data generated later in life. Participants were recruited in part through BabySeq Project sites and those leading genomic screening programs, resulting in our sample being weighted toward professionals with prior exposure to population-scale genomic sequencing, whether through the BabySeq Project or through leading genomic screening programs. This may have produced both more informed and more favorable views of lifelong genomic medicine than would be found among the broader professional community. Genomic screening implementers in particular may have professional or institutional commitments to expanding genomic screening. About 40% of participants came from institutions in two cities, and all adult genetic counselors came from one institution, so these participants may share institutional assumptions and experiences. Nine of 10 PCCs were pediatric clinicians, so adult primary care perspectives, which are relevant to the pediatric-to-adult transition and to adult-onset findings, are largely absent. Group sizes were small and some participants held more than one role, so comparisons between stakeholder groups should be read with caution. While we recruited across multiple professional groups to enable triangulation, we did not include patient or public perspectives; the extent to which professional assessments of patient concerns — around consent, trust, masking, and privacy — reflect patients’ own views remains an open question (we are exploring parents’ views in another study). Participants were predominantly White (36 of the 48 who provided demographic data). Participants were introduced to lifelong genomic medicine as sequencing near birth and querying the data over a lifetime, but the activities this would involve were not specified, so participants may have had different activities in mind. In addition, lifelong genomic medicine does not yet exist as an implemented program: participants were reasoning about a largely hypothetical model, and the challenges they anticipate may differ from those that emerge in practice.

### Conclusions

Professionals across five stakeholder groups saw clinical value in maintaining genomic data across a lifetime, and many expected it to happen. The component pieces are no longer hypothetical: genomic newborn screening is being researched across multiple countries, “sequence once, query often” workflows are already implemented in some health systems, and variant reclassification and *ad hoc* recontact are routine in clinical genetics. Table 3 sets out what already has operational precedent, and in several areas the question is one of scale and funding rather than of invention. Whether all of these activities add enough clinical value to justify lifelong storage and reanalysis is not yet established. Participants also did not identify comprehensive, consistently implemented governance structures that could sustain these practices as a coherent, lifelong intervention, and described responsibility as fragmented, unfunded, or unassigned across nearly every dimension required, from accountability for acting on reclassified variants, to stewardship of data whose significance will change over decades, to maintaining contact with individuals who may not know they were sequenced. Until the evidence on value is clearer, scopes of responsibility are defined, and structural prerequisites are in place, we conclude that lifelong genomic medicine risks proceeding faster than the frameworks needed to make it responsible.

## Supporting information

S3 Table. Coding framework with code frequencies by stakeholder group

## Statements

## Acknowledgements

We would like to acknowledge all our participants, and Ali MacLeod, Mai Ly Burke, Julia Mizzi, Jazmine Harry, Yuka Kato, Layla Horwitz, and Lily Hamilton for their careful work cleaning transcripts.

## Statement of Ethics

This study was performed in accordance with the Declaration of Helsinki. This human study was approved by Mass General Brigham Institutional Review Board, which judged it exempt (approval 2/2/2024, protocol number 2023P002704). Written consent was not required. Before beginning the interview, we confirmed that the prospective participants would like to continue on to participate, having reviewed the information in the fact sheet associated with the study. By continuing they provided implied consent for study participation, as approved by the IRB.

## Author Contributions

ACFL conceptualized the study, acquired funding, conducted all data collection and curation, led the formal analysis, administered the project, created all visualizations, and wrote the original draft. IAH contributed to formal analysis, provided supervision, and conducted validation. RCG acquired funding and provided supervision. All authors contributed to methodology and reviewed and edited the manuscript. All authors read and approved the final version.

## Data Availability Statement

The qualitative nature of the data and the small, specialized professional community from which participants were drawn mean that interview transcripts cannot be shared publicly without risk of participant re-identification, even after de-identification. Participants were assured of confidentiality, and the study was approved under these conditions by the Mass General Brigham IRB. The interview guide, participant fact sheet, and complete coding framework with code frequencies by stakeholder group are provided in the online supplementary material. Requests for additional information about the analytical process may be directed to the corresponding author.

## Online Supplementary Material Captions

**S1 Text. Participant fact sheet.** Information sheet provided to prospective participants prior to enrollment, describing the study purpose, procedures, and confidentiality protections.

**S2 Text. Interview guide.** Semi-structured interview guide used across participant groups. Most components were common across all provider types; sections specific to particular participant groups are indicated.

**S3 Table. Coding framework with code frequencies by stakeholder group.** Complete coding framework showing all codes, their grouping into subthemes and themes, and frequencies by participant group (clinical geneticists, genetic counselors, laboratory personnel, primary care clinicians, genomic screening implementers).

**S4 Checklist. COREQ checklist.** Consolidated Criteria for Reporting Qualitative Research (COREQ) 32-item checklist for interviews and focus groups.

## Mass General Brigham

Study Title: An interview study to help assess the clinical value and feasibility of revealing genetic information at multiple points of the lifespan

Principal Investigator: Robert Green, MD, MPH

Fact Sheet

- This is a Mass General Brigham research study. The purpose of this research is to help assess the value and feasibility of revealing genetic results over the lifespan. This research study is funded by the NIH.
- We are asking you to participate because you are a professional with relevant expertise to help the research team address their research questions. About 64 people will participate in total.
- If you are being emailed directly by the study team, your email was obtained from publicly available information, given your professional role in either a genomic testing lab or a program integrating genomic screening into health systems.
- In this study we will use the platform Zoom to conduct a one-on-one virtual interview. This virtual interview will take 60 minutes and will be recorded. We will ask you questions that draw on your professional expertise; there is no preparation needed on your part.
- You will receive a $100 check for your time. We will need to collect your social security number and address to issue the check. This information will be kept secure and will not be used as study data.
- This interview will be transcribed and analyzed by the study team. We may use direct quotes from participants when presenting our findings.
- The video recordings will only be available to the study team. Your de-identified information may be used or shared with other researchers without your additional informed consent.
- The main risk is a potential loss of privacy and/or confidentiality. We have multiple procedures in place to protect privacy and confidentiality. While we cannot guarantee that the identity of the participants will not be revealed in the event of a breach or other unexpected event, we have precautions in place to keep your information secure. Study data will only be accessible to study staff; your identifiers will be separated from your responses and all data will be coded. We will use the secure systems at Mass General Brigham, endorsed by our Information security team, to secure both your study data and the information provided for issuing the check.
- Participation is voluntary and you can stop at any time.

Anna C F Lewis, DPhil, is the main point of contact for this research study. You may contact her through email at. The PI’s contact information is. If you’d like to speak to someone not involved in this research about your rights as a research subject, or any concerns or complaints you may have about the research, contact the MGB Human Research Committee at (857) 282-1900.

Interview guide for “An interview study to help assess the clinical value and feasibility of revealing genetic information at multiple points of the lifespan”

### For everyone

#### Preamble

Hello, []. Thank you for agreeing to take part in this interview study. My name is Anna Lewis and I’m a research scientist at Brigham and Women’s Hospital division of genetics. My mentor is Dr Robert Green and he is the PI of this study.

As a reminder, we are conducting a research study. The purpose of this interview is to help assess the value and feasibility of revealing genetic results over the lifespan. There are no right or wrong answers to any questions.

I sent you a fact sheet about this study. In brief, this interview will be recorded and then transcribed and analyzed. All identifying information will be removed and all of the analyzed data will remain anonymous. This interview will last about 60 minutes. For participating in this interview, we will send you a $100 check as a thank you for your time. Your participation in this interview is completely voluntary and if at any point you wish to stop the interview, you are welcome to do so. Additionally, if there is a question you would rather not answer, just let me know.

As a thank you for your time, we will send you a $100 check. Per hospital policy, I need to ask you for your SSN in order to issue the check. I’ll do so at the end of the interview; it will be used solely for this purpose, kept secure, and will not be connected to your responses.

Before we get started, do you have any questions about what I have presented, or any questions about the Fact sheet?

Do you consent to be part of this study? Is it okay if I begin recording now?

**Introduction to the Project:** Frame some of the key pieces of background, as well as the motivations and concerns. Explain why they were selected to be interviewed.

**1. Background Questions:** This basic set of open ended questions are designed to get the respondent comfortable sharing and telling us about their work.

- What is the focus of your professional practice?
- What led you to this area?

***For health care providers (Aim 1a)***

## 2a. Reactions to receiving screening information

First ask for examples of where they receive both genetic and (where appropriate) non-genetic examples. For these examples, ask:

- What makes it useful?
- What makes it a burden?
- How does uncertainty factor in?
- Examples of receiving information before they would initiate a risk mitigation strategy?

○ Idea of “patients in waiting”

## 3a. How they view their obligations over time to patients and their parents

- Example of a variant of uncertain significance for a child with a phenotype
- Example of a child with a pathogenic variant but not yet showing signs of having the condition (low penetrance?)

## 4a. Their experiences of longitudinal relationships with patients and their parents

Any experiences of patient-parental divergence of opinion with regards to genetic knowledge?
Any observations of the resources families need access to in order for the value of genetic information to be realized?

## 5a. Attitudes toward when information should be shared

- Are there example genetic conditions where you think it would be optimal to do population screening later in childhood?
- In other words, do you see any clinical value in withholding any later childhood onset information?

## 6a. Their expectations of what patients and parents would want

- What would you anticipate parents would want when it comes to just-in-time information, versus receiving it all at once?
- What about patients themselves, as they reach the age of assent?

**For those with operational roles in generating genomic reports (Aim 1b): not all questions will be relevant to all roles**

## 2b. [Lab personnel] Treatment of sequence information after an initial report is generated

- Current treatment -- does anything happen? For example, reinterpretation of variants?
- Do you envision this changing in the nearterm, and why?

## 3b. [Those embedded in health system] Vision for integration of genomic information into health system

- What is the near term vision for the integration of genomics into the health system?
- [If not in the near term vision] Do you share a medium term vision of using the genome as a resource over the lifespan? Why or why not?

## 4b. How they view their obligations to the individuals who they provide testing for

- Does the lab have any legal or regulatory responsibility to the individuals you provide reports for, after the report is signed out?

○ What about reporting variants that have taken on a new significance?
- Do you think the lab has any *ethical* responsibility in this area?
- Do you think anyone else, perhaps the ordering physician, has any ethical responsibility in this area?

## 5b. [Lab personnel] Whether, and how, their lab currently distinguishes in theory and in practice genetic information from the querying of that information

When a lab generates a variant file, in some sense there is information that is both “there and not there” -- a simple look up could reveal a pathogenic variant, but it won’t necessarily be looked up.

- Does your lab distinguish in theory or in practice genetic information from the querying of that information?

## 6b. Privacy, security and longevity of information

- What do you consider necessary, currently to maintain the privacy, security and longevity of sequence information?
- Do you see any of this changing in the near or mid term?

***For everyone***

## 7. Their views on using the genome as a resource over the lifespan

- Do you think that this vision, of sequencing once and querying many times, is one that the field should be pursuing?

○ Why or why not? Any considerations we haven’t covered? (anticipate a mixture of practical and ethical concerns)
○ Any additional practical considerations that we haven’t touched upon?
○ If ethical concerns haven’t come up explicitly: Do you have any ethical concerns?
○ If equity hasn’t come up: Do you foresee any equity barriers?

## 8. Wrap up

- Is there anyone else you recommend I should talk to about these kinds of questions? Particularly someone who might have a contrasting view to your own?

○ If so, can I send you a recruitment email, for you to forward to this/these individuals, copying me?
- Is there anything else you would like to add?
- Do you have any questions for me?

I am going to stop recording now. Please can I ask you for your SSN? I will store this securely and we will only use it to issue your check. Can I also please confirm what address you would like the check sent to?

Thank you!

## S4 Checklist. COREQ (COnsolidated criteria for REporting Qualitative research) Checklist

Developed from: Tong A, Sainsbury P, Craig J. Consolidated criteria for reporting qualitative research (COREQ): a 32-item checklist for interviews and focus groups. *International Journal for Quality in Health Care*. 2007. Volume 19, Number 6: pp. 349–357.

Manuscript: Sequencing for a Lifetime: Value, Feasibility, and the Governance Gap in Lifelong Genomic Medicine

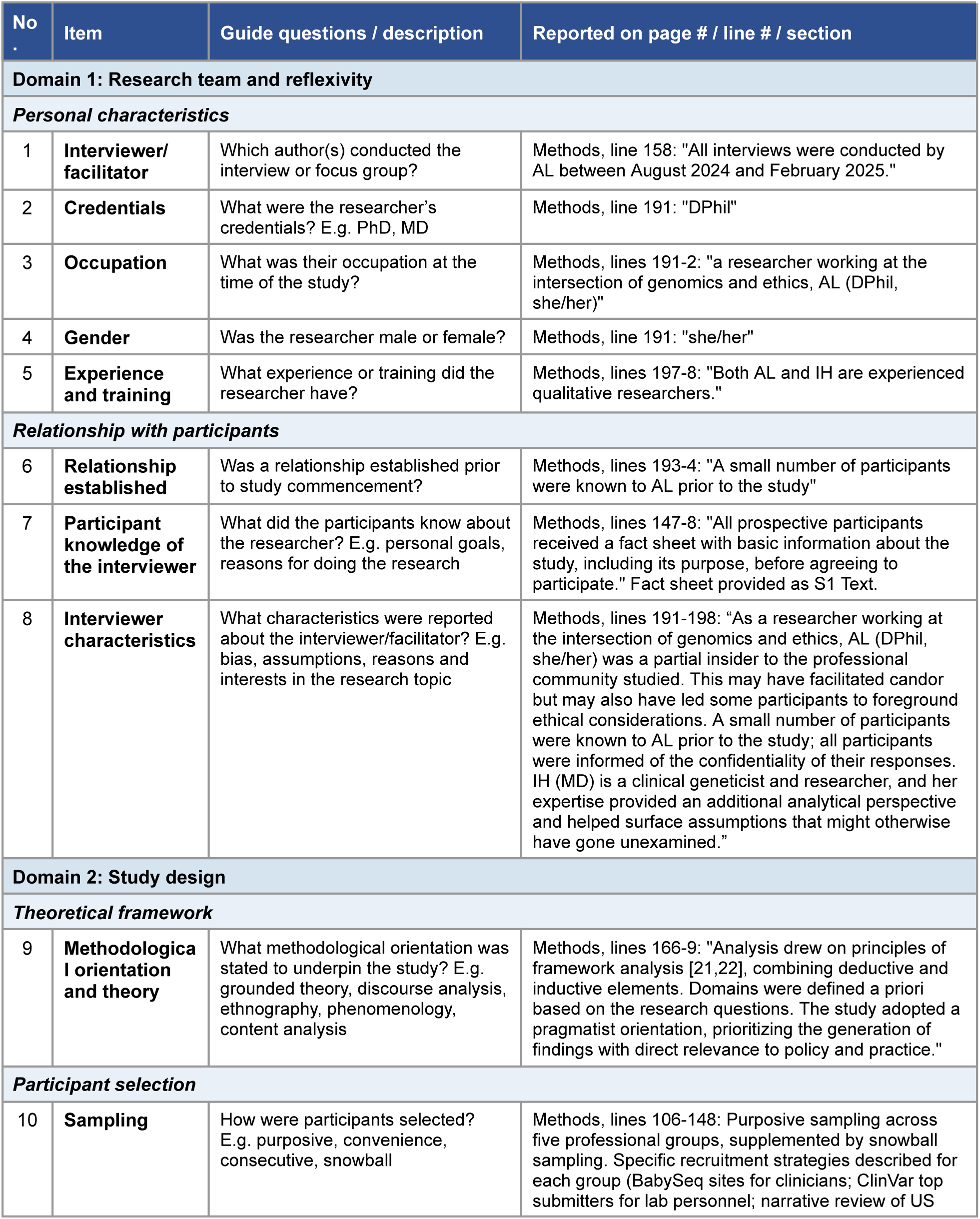
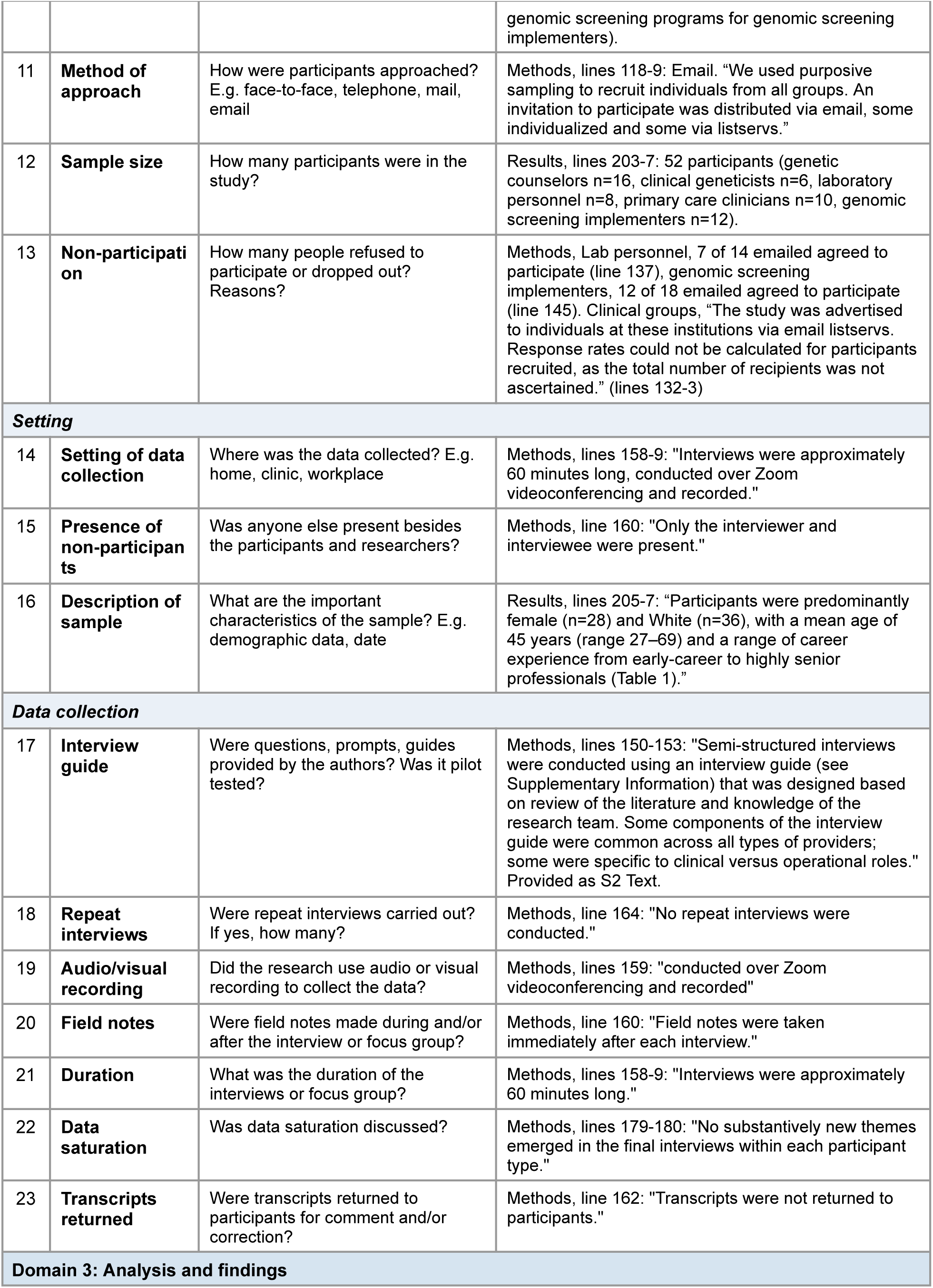
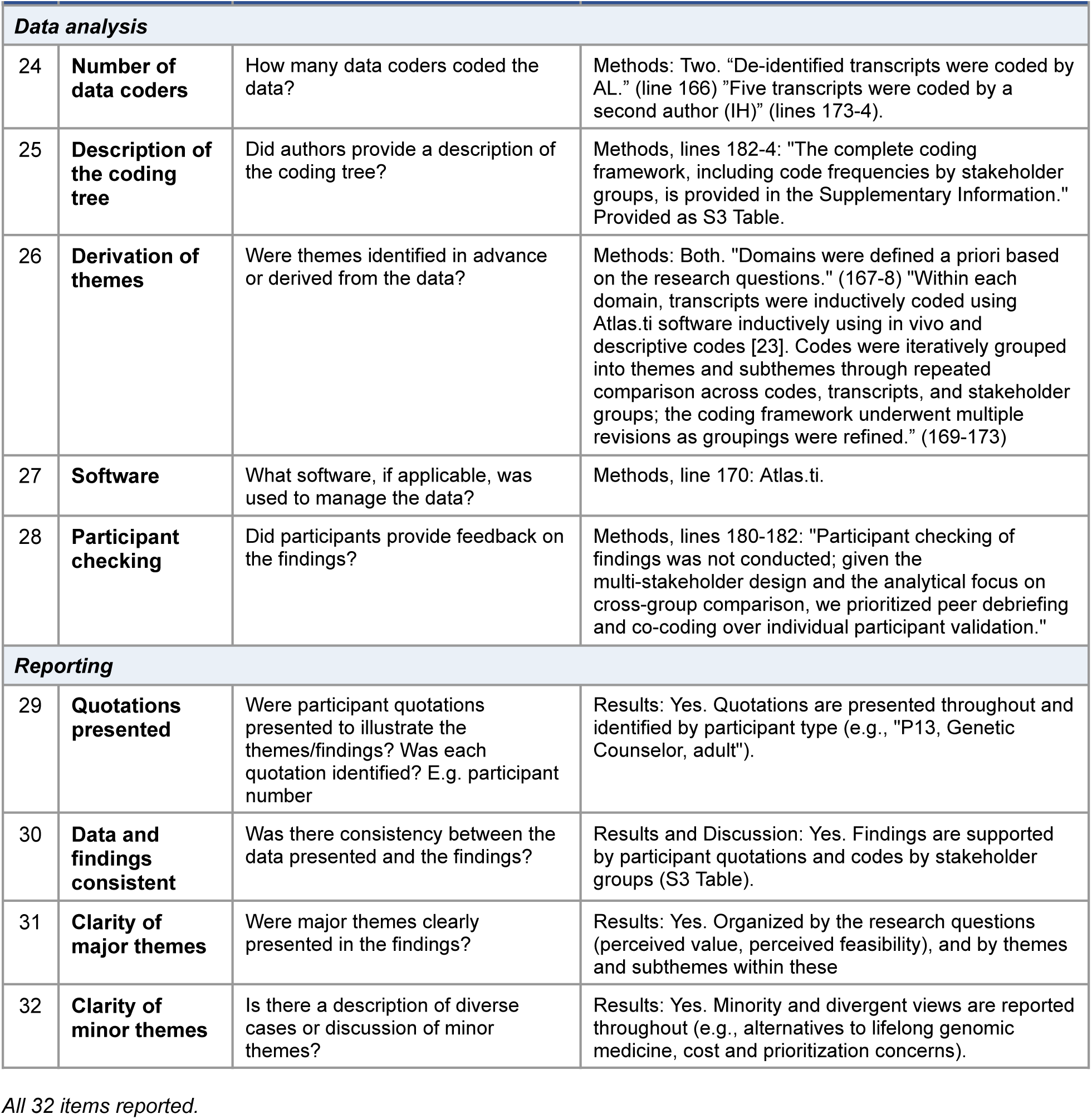

